# Evaluation of Hepatitis B core related antigen (HBcrAg) as a biomarker in cohorts from the United Kingdom and South Africa

**DOI:** 10.1101/2024.07.29.24311156

**Authors:** Louise O Downs, Marion Delphin, Marije van Schalwyk, Susan Hugo, Shiraaz Gabriel, Sheila F Lumley, Elizabeth Waddilove, Tingyan Wang, Catherine de Lara, Arran Babbs, Sue Wareing, Polyxeni Fengou, Monique I Andersson, Richard Glashoff, Jacqueline Martin, M. Azim Ansari, Kosh Agarwal, Geoffrey Dusheiko, Jantjie Taljaard, Wolfgang Preiser, Eleanor Barnes, Gavin Kelly, Ivana Carey, Yusuke Shimakawa, Tongai Maponga, Philippa C Matthews

## Abstract

**Introduction:** Better understanding of hepatitis B virus (HBV) biomarkers is needed. We evaluated Hepatitis B core related antigen (HBcrAg), in the United Kingdom (UK) and South Africa (SA).

**Methods:** We undertook a cross-sectional retrospective observational study of adults with chronic HBV infection from the UK (n=142) and SA (n=211). We recorded clinical and laboratory parameters and quantified HBcrAg. We report HBcrAg distribution, relationship with other biomarkers, and performance in risk stratification based on point of care test (POCT) thresholds.

**Results:** SA and UK cohorts were similar in sex and age (p>0.05), but significantly different in ethnicity, HIV coinfection, HBeAg-positivity and proportion with HBV viral load (VL) >200,000 IU/ml (all p<0.001). More of the untreated SA population had HBcrAg >5.3 log_10_U/ml compared to the UK (33% vs 9% respectively, p<0.0001). HBcrAg ≥4.3 log_10_U/ml (corresponding to a positive POCT) was 100% sensitive and 92% specific for predicting VL >200,000 IU/ml in the UK, but not SA. HBcrAg positively correlated with alanine transferase (ALT) (p<0.001 in UK, p<0.01 in SA), and fibrosis/cirrhosis by APRI score (p = 0.03 in UK and p=0.008 in SA), but not by elastography or FIB-4 scores.

**Discussion:** HBcrAg distribution and relationship with other biomarkers differs between settings. HBcrAg (including POCT) may be a useful proxy for VL, but less so as a marker of disease progression, Its use needs tailoring to represent diverse populations.

## INTRODUCTION

A WHO report published in 2024 estimated 254 million people worldwide are living with chronic hepatitis B infection (CHB), resulting in ∼1.1 million deaths per year as a result of liver disease, including cirrhosis and hepatocellular carcinoma (HCC)^1^. There is a shortage of reliable, accessible and affordable clinical tools to predict outcomes of CHB and its treatment, so new approaches are needed. More precise identification of people at risk of liver complications could inform individualised interventions, such as enhanced surveillance, treatment or preventive interventions, while also providing evidence to underpin resource allocation, clinical services, and development of public health programmes. Evidence that supports these interventions will contribute to progress towards international targets for hepatitis B virus (HBV) elimination as a global public health threat by 2030^2^.

National and international HBV treatment guidelines stratify patients for antiviral therapy with nucleos/tide analogue (NA) drugs based on a combination of serum markers and liver imaging^3,4^. Hepatitis B ‘e’ antigen (HBeAg) positivity, high HBV DNA viral load (VL) and high hepatitis B surface antigen (HBsAg) titres are all associated with progression to cirrhosis and HCC^5–7^. The performance of clinical laboratory liver markers such as alanine aminotransferase (ALT) for predicting liver fibrosis is inconsistent, in some studies this marker had high sensitivity for predicting liver fibrosis^8^, whilst in others it was a very poor predictor of any fibrosis grade^9^.

All current serum biomarkers are imperfect predictors of outcome, and not universally available across all settings due to infrastructure and resource constraints. The landscape is changing as new WHO guidelines on the management of HBV released in 2024 have relaxed criteria for treatment^10^, such that a wider population become treatment eligible, based on assessment with a less complex algorithm. This shift calls for cheaper and more widely accessible tools for monitoring, and predictors of outcomes that can be applied to those both on and off NA treatment. Meanwhile, as search for cure therapies continues, there is also a need to develop biomarkers that can support a mechanistic understanding of liver pathology and approaches to monitoring^11^, and also increasing focus on biomarkers which might help support safe treatment cessation^12^.

NA agents control HBV replication by inhibiting the viral reverse transcriptase (RT) enzyme, thus abrogating production of new HBV DNA genomes and secretion of new infectious virions. However, NA treatments have no direct influence on the intracellular viral reservoir in the form of covalently closed circular (ccc)DNA, and rebound viraemia therefore typically occurs if treatment is stopped. Monitoring of cccDNA levels currently requires invasive investigations and is not possible in routine clinical practice. Hepatitis B core related antigen (HBcrAg) is a circulating serum biomarker that combines HBeAg, hepatitis B core antigen (HBcAg), and a small 22kd protein called p22cr^13,14^. HBcrAg is a surrogate marker of active viral cccDNA transcriptional activity, shown through comparison of serum HBcrAg levels and cccDNA quantification in liver biopsy^11,13,15^, and also correlates with peripheral HBV VL and HBeAg status.^16,17^ HBcrAg has been associated with non-invasive scores of liver inflammation or fibrosis (Aspartate aminotransferase (AST) to Platelets Ratio Index (APRI) and Fibrosis-4 (FIB-4) score), Alanine Aminotransferase (ALT) levels^18,19^, hepatic necroinflammation^15,16^, HCC^20^, and liver flares after treatment cessation^21^. One study of >400 Asian treatment-naïve patients reports significant changes in HBcrAg associated with different disease ‘stages’^22^. Over 10 years of follow up, HBcrAg was a better predictor of HCC than HBV DNA^23^.

HBcrAg could also be of utility as a biomarker of viral reservoir or replication when HBV VL is low or tools for VL quantification are not available, as in much of the world. In one study after 7 years of follow up, over 50% of HBeAg-negative patients (primarily with undetectable VL) still had detectable HBcrAg^24^. WHO guidelines highlight HBcrAg assays as a potential approach for improving risk stratification^10^, although these tools remain in the research domain as there are no commercially available assays. To date, there are limited data on HBcrAg representing diverse populations, with heterogeneity of results, especially depending on HBeAg status. Most existing studies investigating HBcrAg are in Asian populations (therefore representing HBV genotypes B and C), or in Europe, thus neglecting representation of individuals of African descent^25,26^. To date, there is only one study of HBcrAg in an African population (conducted through the PROLIFICA cohort in the Gambia), which showed that HBcrAg could predict clinically important HBV VL thresholds^27^.

To move towards more accessible HBcrAg measurement, a point of care test (POCT) has been developed and validated in the PROLIFICA cohort^28^ to identify highly viremic patients, with an estimated threshold for positivity at 4.3 logIU/mL. This could enhance timely clinical assessment, support early treatment decisions, and promote linkage to care and continuity of follow-up^28,29^.

To add to the body of available evidence for the use of HBcrAg as a biomarker, we studied two cohorts recruited in distinct geographical locations, the United Kingdom (UK) and South Africa (SA) (**Figure 1**). We set out to generate a cross-sectional analysis of the variation of HBcrAg within and between cohorts, and to describe its relationship with host characteristics and routine baseline clinical biomarkers (HBeAg and VL). We then undertook analysis to determine how HBcrAg might have utility in predicting: (a) laboratory markers of liver inflammation (ALT); (b) clinical liver disease progression, including fibrosis and cirrhosis, assessed using APRI, FIB-4 and elastography scores; and (c) clinically relevant VL thresholds (such as those established for assessing eligibility for perinatal prophylaxis). On this basis, we aim to contribute to the dialogue about the real-world use of HBcrAg to support delivery of interventions for HBV surveillance, treatment and prevention.

**Figure 1:**
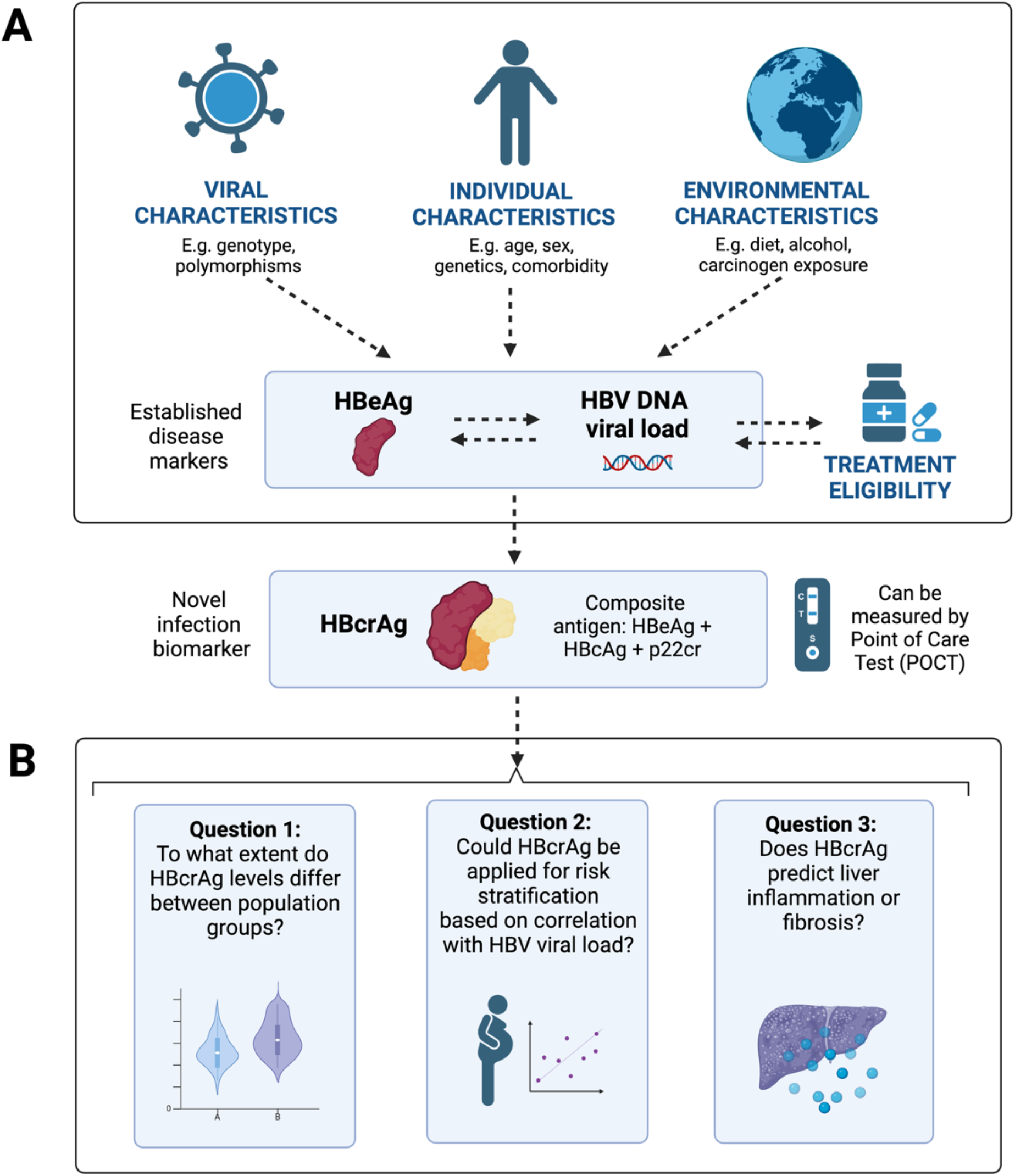
Schematic to show the known and potential relationships between HBcrAg and other viral, host and environmental factors. (A) External factors which may influence HBcrAg; (B) Research questions to determine the potential utility of HBcrAg measurement. Figure created in Biorender with a licence to publish.

## Methods

### Study recruitment and collection of samples and data

We undertook a cross-sectional retrospective observational study, using serum samples obtained from adult patients with CHB enrolled at Oxford University Hospitals (OUH) NHS Foundation Trust (UK, n=142), and from the ‘OXSA-Hep’ Cohort in Cape Town and Bloemfontein, South Africa (SA; n=211). Study settings and recruitment have been previously described for the UK^30,31^ and SA^32–34^. Ethics approval was provided by Oxford Research Ethics Committee A (reference 09/H0604/20 and N17/01/013), University of Oxford Tropical Research Ethics Committee (ref. OXTREC 01–18), Stellenbosch University Health Research Ethics Committee (HREC ref. N17/01/013), and the University of the Free State Research Ethics Committee (HSREC ref. 203/2016). All participants provided written informed consent.

Samples were transported to the research laboratory within 2-3 hours of collection, spun to separate serum, and frozen in aliquots at -80°C. We recorded patient demographics, ALT, presence of HIV and elastography scores from routine clinical and electronic patient records. The serum sample collection date was designated ‘baseline’, and metadata including ALT, elastography scores and markers for calculation of non-invasive liver scores were collected as close to baseline as possible, with a limit of 12 months either side of this date.

We calculated laboratory fibrosis scores using the following formulae:

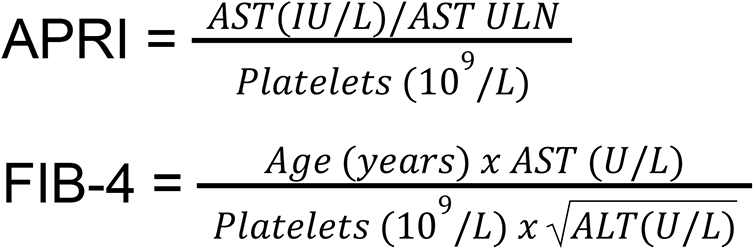

For the purposes of this calculation, AST upper limit of normal (ULN) was set as 40 U/L.

### Quantification of standard serum HBV parameters

Platforms used to measure clinical laboratory biomarkers (HBeAg, VL, ALT, AST, Platelet count) in the Oxford cohort (UK) and Stellenbosch/Bloemfontein cohort (SA) are listed in Suppl. Table 1. Assays were undertaken on validated clinical diagnostic platforms according to the manufacturers’ instructions.

### HBcrAg Measurement

Quantitative levels of HBcrAg were determined using the Lumipulse G HBcrAg chemiluminescent immunoassay (CLEIA) (Fujirebio, Tokyo, Japan) at King’s College Hospital NHS Foundation Trust, London, UK according to manufacturer’s instructions. The assay has a linear measurement range of 3-7 log_10_ U/mL, using 3 log_10_ U/ml as the lower limit of detection (as recommended by the manufacturer). Samples generating HBcrAg measurements >7 log_10_IU/ml were diluted (1:5, 1:100 or 1:1000 in Tris Buffered Saline) before repeat testing.

### Thresholds for biomarkers and liver elastography

We used thresholds for analysis based on existing precedents and 2024 WHO guidelines^10^, including HBV VL >2000 IU/mL, >20,000 IU/mL and >200,000 IU/mL, which have been established clinically to determine eligibility for antiviral treatment and/or prophylaxis,^35^ and ALT thresholds for upper limit of normal (ULN) of 19 IU/L in females and 30 IU/L in males. We defined the presence of liver fibrosis based on FIB-4 ≥1.45, APRI >0.5 or elastography score >7kPa, while cirrhosis was defined as FIB-4 ≥3.25, APRI >1 or elastography score ≥12.5 kPa^10^. We considered HBcrAg thresholds based on those previously determined as associated with relevant HBV VL thresholds^17^, where HBcrAg 3.6 log_10_U/ml, 4.5 log_10_U/ml and 5.3 log_10_U/ml were predictive of HBV VL >2000 IU/mL, >20,000 IU/mL and >200,000 IU/mL, respectively^10^. Previous data from the Gambia established the HBcrAg POCT to have a HBcrAg laboratory cut-off of approximately ≥4.3 (log_10_U/ml)^28^. Whilst acknowledging we did not undertake POCT in this population, we applied this as a binary cut-off to analyse the relationship with liver health markers in each cohort. We also calculated the sensitivity and specificity of our estimated positive HBcrAg POCT in predicting clinically significant thresholds that are applied to inform intervention.

### Data Analysis and Statistical Methods

We used R version 4.2.0 and GraphPad Prism version 10.1.0. We did not pool data for SA and UK cohorts, but analysed each independently, recognising that the genetic, clinical, epidemiological, and environmental landscape of our two settings is not comparable, with multiple unmeasured influences which could contribute to liver disease and infection outcomes. Instead, we compared the findings between the two settings to review whether similar assumptions and uses of HBcrAg can be applied uniformly in different geographical populations. We focussed primarily on those not taking NA therapy, to assess the use of HBcrAg in making treatment decisions. The overall plan for analysis of this data including the questions we are addressing is shown in **Figure 1**.

***(i) Descriptive statistics***

We produced descriptive statistics for each cohort including median and interquartile ranges for patient demographics and HBV biomarkers. We generated comparative statistics within each cohort using Pearson’s, Chi-squared test and Wilcoxon rank sum test.

We also undertook analysis of two distinct subpopulations within each cohort, based on a specific rationale about the potential utility of HBcrAg in each.

> **HBeAg-negative**: In individuals who are HBeAg-positive, HBcrAg levels are likely to be dominated by HBeAg and independent measurement of HBcrAg as a biomarker may therefore not add any additional value. However, we hypothesised that HBcrAg may provide useful additional characterisation within the HBeAg-negative subpopulations.
> **Those not taking NA therapy:** For analyses investigating correlations between HBV VL and liver fibrosis markers with HBcrAg, we included only those people currently off NA therapy. This removes some confounders including varying treatment duration and HIV status, and provides the most useful interpretation for practical application of these findings in the real world. We do note however, that some people may have previously taken treatment so may not be completely treatment naive.

***(ii) Univariable analysis of relationship between HBcrAg and other parameters***

Within each cohort we described the distribution of HBcrAg and explored its relationship with individual demographic factors (age, sex), routine laboratory biomarkers (HBeAg, HBV VL and ALT) and liver fibrosis/cirrhosis scores (elastography, FIB-4 and APRI). When assessing VL and liver fibrosis/cirrhosis scores, we included only those off NA therapy.

***(iii) Sensitivity and specificity with which HBcrAg predicts clinically important HBV VL levels***

In those currently not receiving antiviral therapy, we assessed the sensitivity and specificity of previously defined HBcrAg levels (3.6, 4.5 and 5.3 log_10_U/ml)^17^ and the published laboratory threshold of the POCT (4.3 log_10_U/ml)^28^ to predict clinically important VL thresholds (2000, 20,000 and 200,000 IU/ml respectively), with particular focus on HBcrAg as a risk-stratification tool in a clinical setting to determine eligibility for perinatal NA prophylaxis.

***(iv) Focused multivariable analysis***

We accounted for age, sex, HBeAg status, treatment status and HBV VL in a multivariable model to investigate the extent to which HBcrAg could predict the presence of liver inflammation (based on ALT), fibrosis and cirrhosis (based on elastography, FIB-4 and APRI scores)^36^. For each measure of liver outcome, we independently fitted a binomial generalised linear model with those covariates as main effects along with HBcrAg, cohort and their interaction, and binary liver outcome as the dependent variable. The strength (log-odds ratio) and significance of the association between HBcrAg and liver outcome (stratified by cohort and compared between cohorts) were calculated using bias-reduced score by the `brgrlm` R package. We used complete cases only - patterns of missingness therefore varying between the different measures of outcome.

## Results

### Clinical and laboratory phenotypes of HBV infection in cohorts in the UK and SA

We generated HBcrAg measurements from 353 samples (142 from the UK and 211 from SA); (**Table 1**). Sex and age were comparable between cohorts (p>0.05). However, as anticipated, the cohorts differed in other important ways according to the setting: 68% of SA participants were black African, in contrast to more diverse ethnicities among participants recruited in the UK (p <0.001). Compared to the UK, a higher proportion of the SA population were living with HIV coinfection, were HBeAg positive and had HBV VL >200,000 IU/ml (absolute numbers shown in Table 1; all comparisons p<0.001), although median HBV VL did not significantly differ between cohorts (p=0.3). Reflecting these differences (particularly based on HBeAg-positivity, and HIV coinfection), a higher proportion of People Living With HBV (PLWHB) were receiving antiviral treatment in SA (129/197, 65%) compared to the UK (42/137, 35%); (p <0.001). Based on ALT and APRI measurement, liver inflammation was not significantly different between the UK and SA cohorts (p=0.4 for both). However, FIB-4 and elastography scores were both significantly increased in the SA population compared to the UK (p=0.043 and p=0.025 respectively).

**Table 1:**
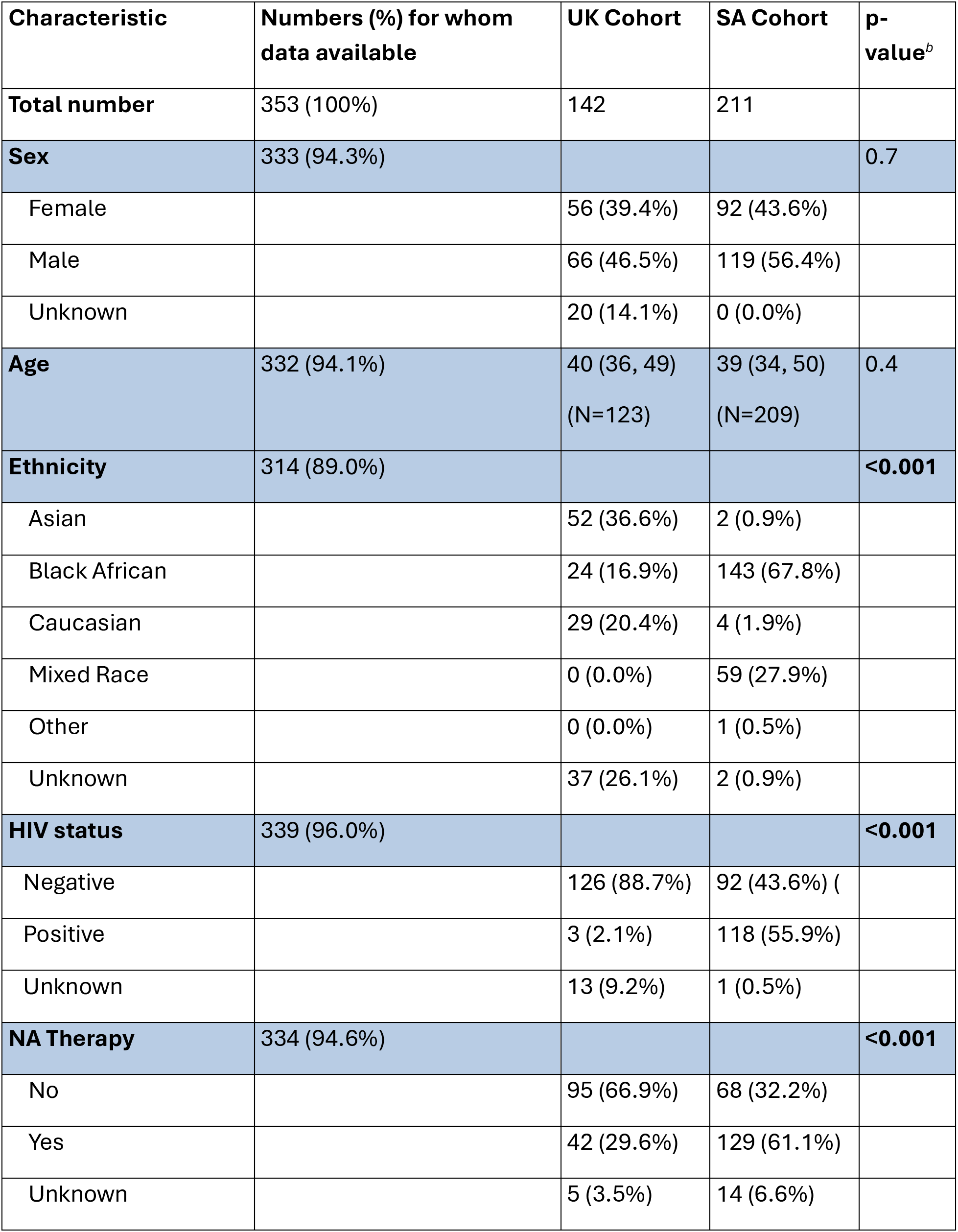
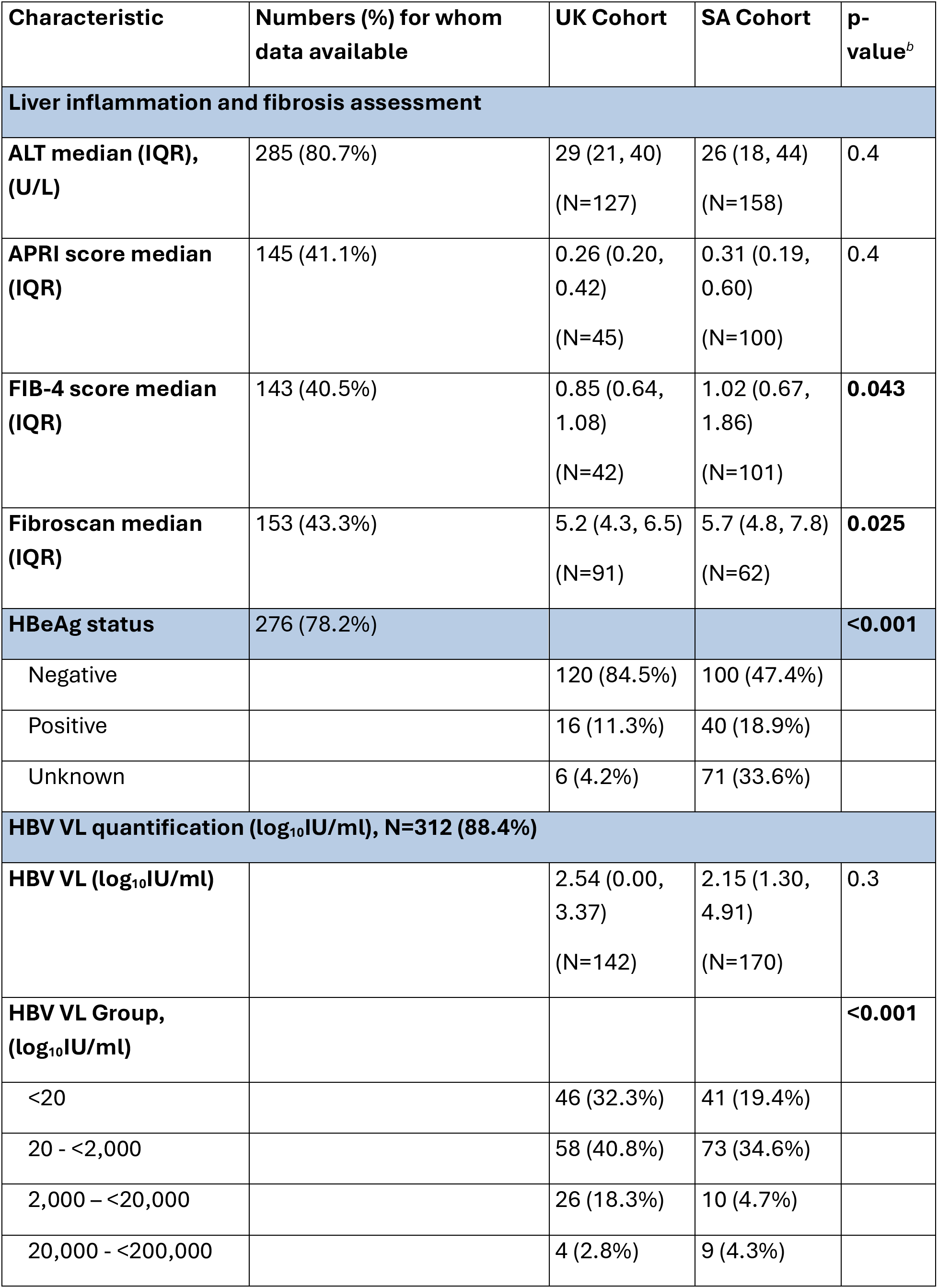
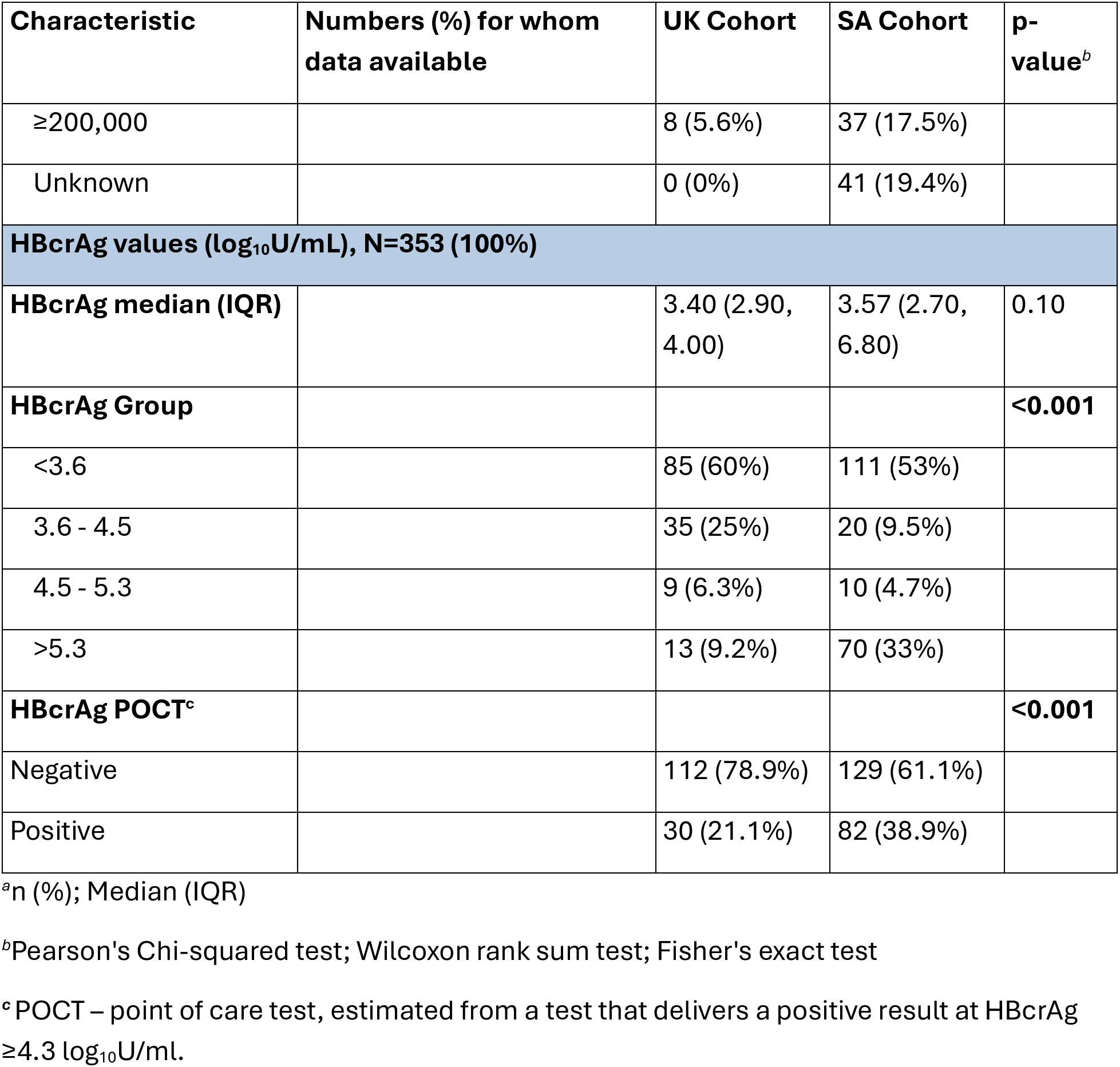
Characteristics of two cohorts of adults with chronic hepatitis B virus infection based in South Africa (SA) and the United Kingdom (UK).

These findings, in addition to differences in environmental exposures, host and viral genetics, clinical, biological and treatment landscapes between settings, underpin our subsequent analysis approach, in which each cohort is assessed independently.

### Distribution of HBcrAg levels in two different settings

The distribution of HBcrAg varied between UK and SA populations (**Table 1, Figures 2A and 2B**), and a greater proportion of those in the SA cohort had HBcrAg ≥5.3 log_10_U/ml (70/211 (33.2%) in SA vs 13/142 (9.1%) in UK, p<0.001, **Table 1**). When estimating the HBcrAg POCT results (threshold = 4.3 log_10_U/mL), 30/143 (21.1%) of the UK population would test POCT positive compared to 82/211 (38.9%) of the SA population (p=0.0005, **Table 1 and Figures 2A and 2B**). However, despite these differences, median HBcrAg was comparable between the UK and SA populations (3.40 log_10_U/ml vs 3.57 log_10_U/ml respectively, p=0.1, **Table 1**). On univariable analysis, HBcrAg was not associated with sex (p>0.05), but had a significant negative association with age in both cohorts (p=0.01 in UK and p=0.004 in SA, **Table 2**).

**Figure 2:**
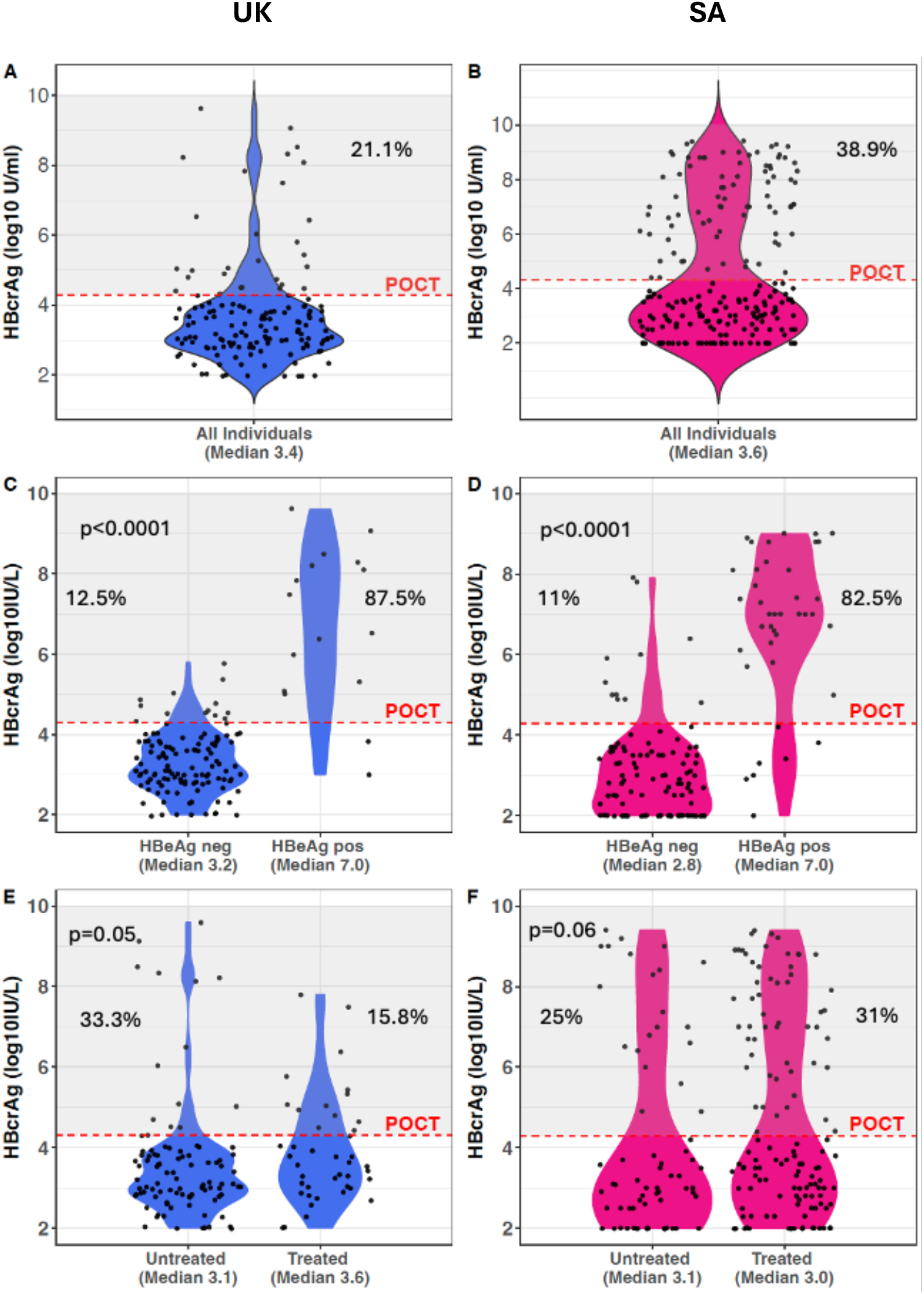
HBcrAg distribution in adults living with chronic hepatitis B (CHB) in South Africa (SA) and the United Kingdom (UK): A: UK cohort, all patients, B: SA cohort, all patients, C: UK cohort split by HBeAg status, D: SA cohort split by HBeAg status, E: UK cohort split by treatment status, F: SA cohort split by treatment status. p-values present Mann-Whitney-U test for significance using quantitative CrAg data. Grey areas represent the population who would be identified by a positive POCT (HBcrAg≥4.3 log U/ml); indicated next to each violin is the percentage of tests which meet this threshold.

**Table 2:**
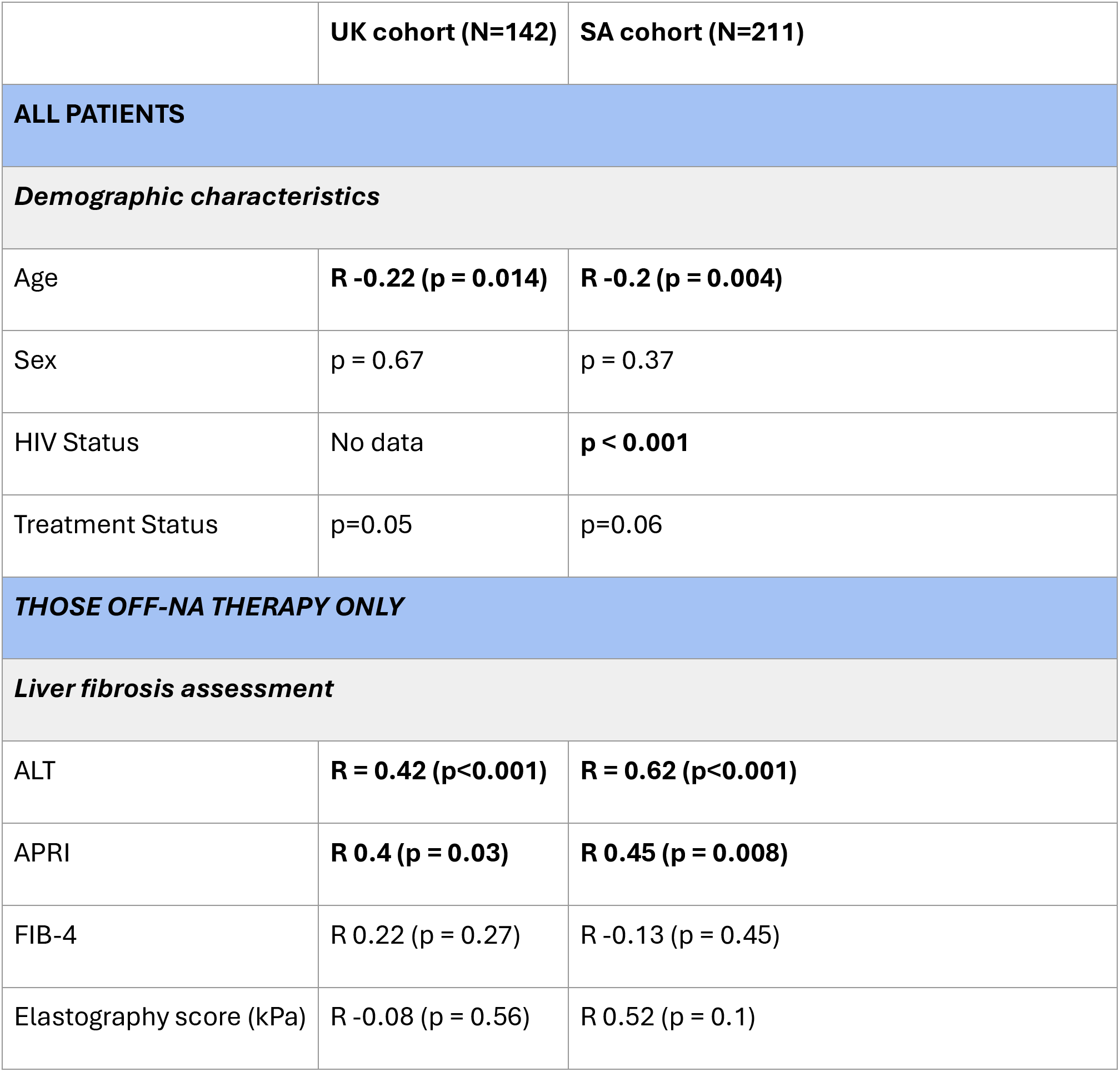
Correlation between HBcrAg and demographic, laboratory and imaging characteristics of adults living with chronic HBV infection in the United Kingdom (UK) and South Africa (SA). Pearson’s correlation coe/icient (R) is shown for continuous variables, Wilcoxon rank sum test is used to compare categorical variables.

As expected, in both cohorts, HBcrAg significantly differed between HBeAg-positive and -negative groups (p<0.001), with 87.5% of HBeAg positive samples meeting the POCT threshold in the UK cohort, and 82.5% in SA (**Figures 2C and 2D**). Median HBcrAg was significantly higher in those living with HIV co-infection compared to HIV-negative participants in SA (3.9 vs 3.3 U/ml, p <0.001; **Table 2**). As HIV infection was infrequent in the UK cohort, we could not investigate any influence of HIV coinfection. HBcrAg was not influenced by treatment status in either cohort (**Figures 2E and 2F**).

### Risk stratification using HBcrAg based on correlation with HBV VL

Considering only untreated individuals, there was a significant positive correlation between HBcrAg and VL in both the UK and SA cohorts (R=0.68 and 0.8 respectively, p<0.001 for both; **Figures 3A and 3B**). However, in the UK cohort, this correlation was lost when considering only the HBeAg negative population (**Suppl Figures 1A** **and** **1B**). In the SA cohort, median HBcrAg was significantly lower in those with undetectable vs detectable VL (3.1 vs 3.8 log_10_U/ml, p<0.001, **Suppl Figures 1C** **and** **1D**).

**Figure 3:**
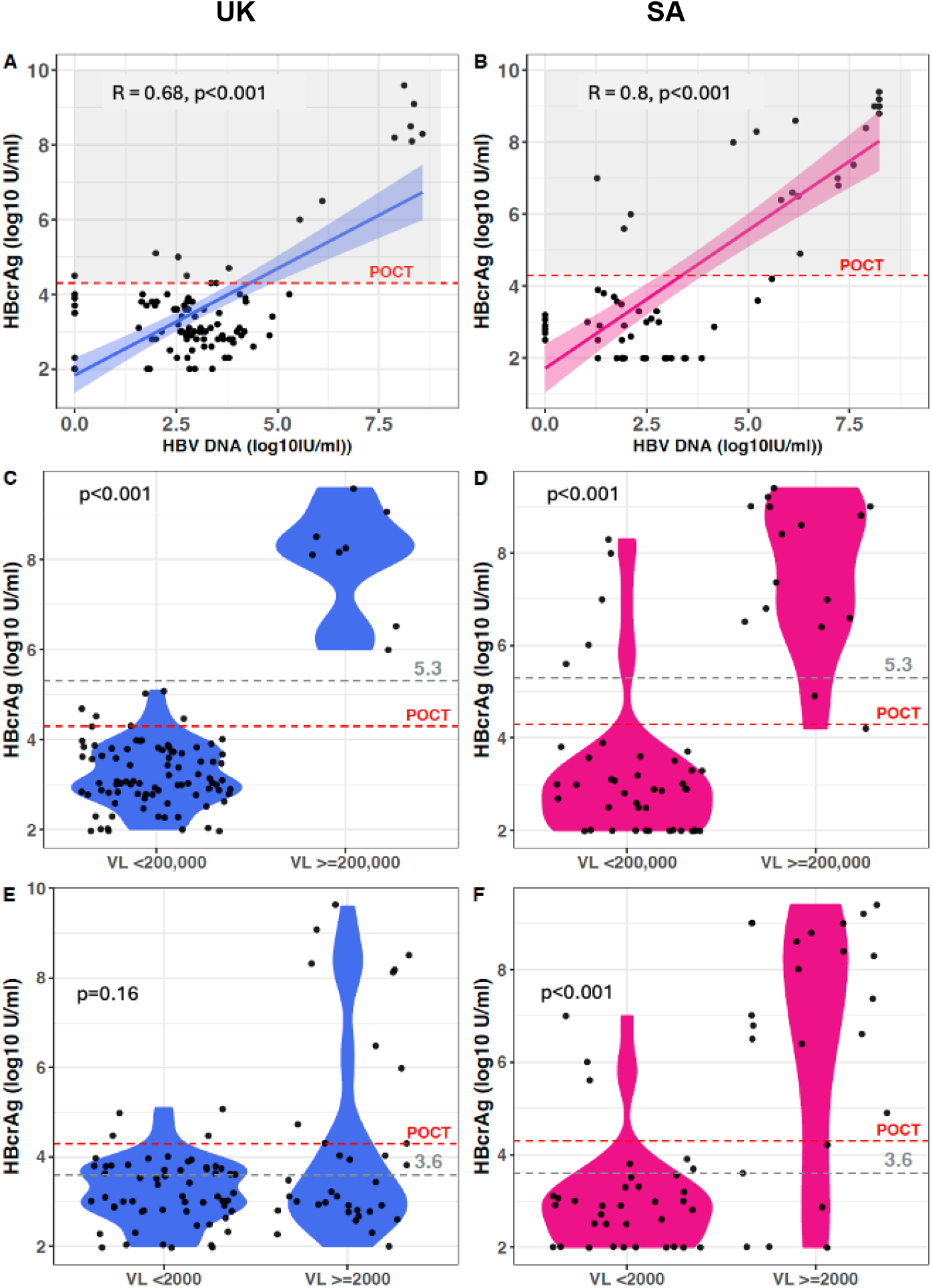
Relationship between HBcrAg and HBV viral load (VL) in adults living with chronic hepatitis B (CHB) in South Africa (SA) and the United Kingdom (UK) not taking NA therapy: A: UK all patients off NA therapy, B: SA all patients off NA therapy, C: UK cohort split by HBV VL <200,000 IU/ml vs ≥200,000 IU/ml, D: SA cohort split by HBV VL <200,000 IU/ml vs ≥200,000 IU/ml, E: UK cohort split by HBV VL <2000 IU/ml vs ≥2000 IU/ml, F: SA cohort split by HBV VL <2000 IU/ml vs >2000 IU/ml. For panels A and B Peason’s correlation is shown, panels C-F, Mann-Whitney-U test for significance is shown. Dotted red lines represent the HBcrAg POCT threshold (HBcrAg ≥4.3 logU/ml) with the shaded grey area in A and B showing all those points that would test positive; in panels C-F dotted grey lines represent the relevant HBcrAg threshold.

When considering the use of HBcrAg in informing treatment decisions, we evaluated its ability to identify those with VL >2000 IU/ml and >200,000 IU/ml (thresholds based on the 2024 WHO guidelines)^10^. HBcrAg ≥5.3 log_10_ U/ml was 100% sensitive and specific at identifying those with VL >200,000 IU/ml in the UK population but performed less well in the SA population (where it was 88% sensitive and specific) (**Table 3**). There was a statistically significant difference in median HBcrAg between those with VL ≥200,000 and <200,000 in both cohorts, p<0.001 (**Table 3, figures 3C and 3D**, **Suppl Figure 2**).

**Table 3:**
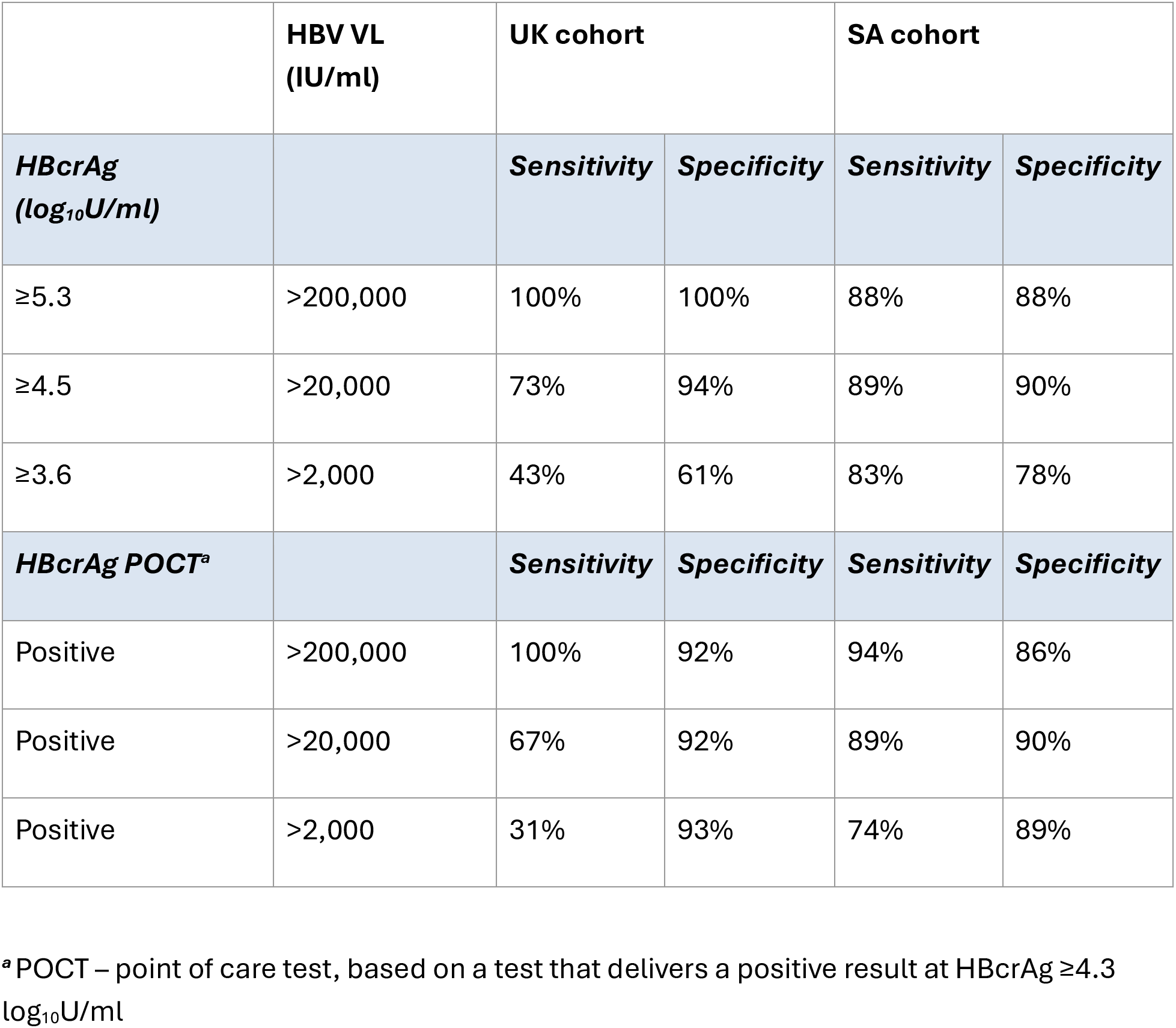
Sensitivity and specificity of HBcrAg levels as predictors of clinically significant HBV viral load (VL) levels in cohorts in the United Kingdom (UK) and South Africa (SA) in untreated individuals.

In both cohorts, the performance of lower HBcrAg levels in predicting defined lower HBV VL thresholds generally declined, particularly in the UK setting, with sensitivity for detecting VL 20,000 IU/ml of 73% in the UK and 89% in SA, and for VL 2,000 IU/ml 43% in the UK and 83% in SA (**Table 3**). In SA there was still a significant difference in HBcrAg between those with a VL >2000 IU/ml and <2000 IU/ml (p<0.001, **Figure 3F**), however this was not the case in the UK where there was no difference (**Figure 3E**).

The reported HBcrAg POCT threshold of 4.3 log_10_ U/ml could not accurately identify those with a VL >2000 IU/ml in either the UK or SA (sensitivity 31% and 74% respectively, **Table 3**), although a greater proportion of those with VL >2000 IU/ml had an HBcrAg above this threshold in both populations (**Figure 3E and F**). In contrast, mothers with HBV VL >200,000 IU/ml at highest risk of vertical transmission could be reliably identified by the POCT threshold in the UK population (100% sensitive and 92% specific), but less so in the SA population (94% sensitive and 86% specific) (**Table 3, Figure 3E and 3F**, **Suppl Figure 2)**.

### Does HBcrAg predict liver inflammation or fibrosis?

In the untreated population, we observed a significant positive correlation between HBcrAg and both ALT and APRI scores in UK and SA populations (ALT: R=0.42 (UK), and 0.62 (SA), p<0.001 for both, APRI: R=0.4, p=0.03 (UK) and R=0.45, p = 0.008 (SA), **table 2**). However, there was no significant correlation between HBcrAg and liver stiffness scores assessed by elastography or FIB-4 in either cohort (**Table 2**). We considered the performance of HBcrAg POCT as a predictor of liver inflammation and fibrosis. Median ALT and APRI scores were significantly higher in those with HBcrAg above this threshold in both populations (p<0.05 for all, **Figures 4C - 4F**), whereas FIB-4 and elastography scores did not significantly differ in either population (**Figures 4G and 4H**).

**Figure 4:**
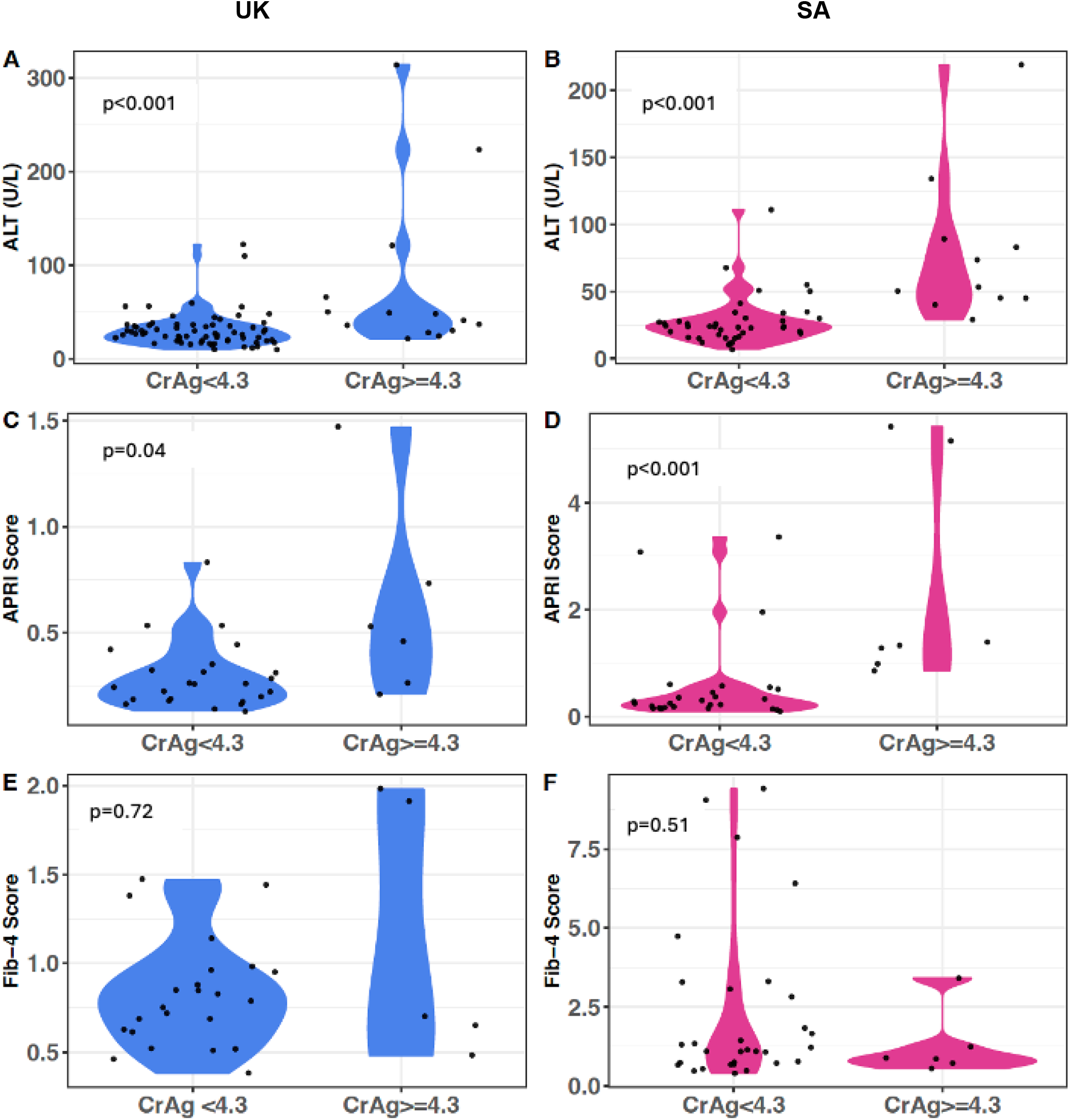
Relationship between HBcrAg POCT threshold of 4.3 log_10_U/ml and liver disease markers in adults with chronic untreated HBV infection. A: Between HBcrAg and ALT in UK cohort; B: Between HBcrAg and ALT in SA cohort; C: Between HBcrAg and APRI in UK cohort; D: Between HBcrAg and ALT in SA cohort; E: Between HBcrAg and FIB-4 score in UK cohort; and F: Between HBcrAg and FIB-4 score in SA cohort. Mann-Whitney-U test for significance is shown.

We applied a multivariable model analysis to determine the magnitude of association of HBcrAg with different liver health indicators across our study populations. This model accounted for covariates identified as potential confounders or predictors of outcome in univariable analysis, including age, sex, HBeAg status, treatment status and HBV VL. The odds of liver outcomes were neither significantly correlated with HBcrAg, nor were they different between the cohorts (**Suppl Table 2**).

## DISCUSSION

### Summary of rationale and findings

We investigated the extent to which HBcrAg could improve clinical assessment (especially if made accessible as a POCT), in two geographically distinct populations, evaluating its relationship with demographic and clinical parameters, especially those that are relevant to treatment stratification.

The only previous study on HBcrAg in Africa (in the Gambia) reports a median of 4.0 log_10_U/mL^27^, which is comparable to that in our SA cohort. However, only 13% of the Gambia population was HBeAg positive, compared to 11.3% in the UK cohort and 18.9% in our SA cohort, which could be related to differences in predominant genotypes. As expected, HBcrAg significantly positively correlated with HBeAg status and with HBV VL in both our study populations; this correlation persisted on excluding the HBeAg positive population in SA but not in the UK, in line with previous observations from the Gambia^27^.

### Clinical utility of HBcrAg in risk assessment

Our observations illustrate the persistence of HBcrAg as a quantifiable biomarker even when VL is low or undetectable. Given the overlap in proteins measured by testing for HBeAg and HBcrAg, the latter could be considered as a more sensitive assay that could supersede conventional HBeAg testing. Elastography score and ALT were previously correlated with HBcrAg levels in untreated patients in the Gambian study^27^; this observation was recapitulated in both our study settings. We also found a correlation between HBcrAg and APRI, although HBcrAg did not correlate with liver fibrosis/cirrhosis based on FIB-4 or elastography scores. While we did not directly evaluate POCT in this study, by applying the equivalent quantitative HBcrAg threshold of 4.3 log_10_ U/ml we demonstrate that POCT could potentially identify those with significant liver disease based on ALT and APRI. The POCT would also be highly sensitive at identifying those eligible for perinatal antiviral prophylaxis in the UK population, but this is less true in SA. These results demonstrate that HBcrAg POCT needs to be evaluated in distinct populations to build a use case, and different population thresholds may be needed.

### Resource impact of HBcrAg

Currently, HBcrAg CLEIA is not available in most settings and comes at a high cost, and running serial dilutions added to the cost of our laboratory assessment of samples with HBcrAg above the upper limit of quantification (although this refined quantification of high levels may not be necessary if the assay were to be deployed in more routine practice). The HBcrAg POCT is currently only available for research purposes, but given our above results, it may have promise for clinical use when other methods of stratification are unavailable. Costs of procuring the POCT must be balanced against cost and lack of availability of other more established monitoring tools that require laboratory facilities, and striking a balance between ongoing investment to develop and evaluate new tools, while avoiding unnecessary cost and complexity. The POCT can be stored at room temperature, does not require specialist equipment and can be done in the field. As turn-around times for diagnosis are often days (and may be weeks in some settings), POCT also adds the benefit of real-time evaluation (turnaround: 45 minutes) leading to prompt advice regarding treatment. A study that reported on screening of migrant populations in Belgium showed that the HBsAg POCT vs venepuncture testing led to a dramatic increase in linkage to care (from 34% to 86%)^29^. Hence, whilst we still need to refine the utility of HBcrAg, there is emerging evidence that POCT can be associated with improved linkage to care.

### Limitations of this study

We measured HBcrAg in modestly-sized cohorts, with significant missingness for some parameters (e.g. fibrosis scores only computed for <50% of individuals) and only on a cross-sectional basis. We have identified some important differences between our study populations in different geographical settings (demographics, HIV coinfection, treatment status), but there are also many unmeasured factors (differences in viral genotype, host genetics, comorbidity, treatment duration, environmental exposures, immune activation, microbiome) that are inter-related and may influence host and viral biomarkers and outcomes of infection. There are some discrepancies between our findings and existing literature^15,37,38^. Differences between our two cohorts, and between this study and others, may be accounted for by (i) methodology used for fibrosis assessment (e.g. histological METAVIR analysis vs. circulating marker scores), (ii) location of study populations and (iii) disease stages, including HBeAg status.

We used the laboratory threshold of 4.3 log_10_U/ml as a substitute for a positive HBcrAg POCT, however we did not actually do the POCT in this group. There is the possibility that the threshold would differ in these geographical locations due to differences in HBeAg positivity or HBV genotypes, although when trialled in the Gambia, there was no difference in the laboratory threshold between genotypes A and E.

Whilst large, longitudinal studies correlating HBcrAg with liver outcome have been done in other populations, this data is missing for Africa where studies on HBcrAg are few, and primarily cross sectional such as this one. Whilst these data are useful to understand the broad utility of HBcrAg and its value in different populations, data evaluating HBcrAg over time will primarily inform its utility in low resource settings, for example whether it can detect changes in liver fibrosis during NA therapy and how closely it can be used where other markers are not available.

## Conclusions

Based on this study, the distribution of HBcrAg levels differs between populations. In our cohorts, HBcrAg does not have a consistent role in predicting liver inflammation or fibrosis, however, HBcrAg correlates with VL, ALT and APRI, and testing may have a role in stratification of interventions for PMTCT, particularly if an affordable POCT becomes available. More data are needed to represent different population settings, and to determine the cost-benefit of introducing new testing at a time when global resource focus is on scale-up of diagnosis and treatment.

## Supporting information

Platforms used to measure routine clinical laboratory biomarkers in Oxford cohort (UK) and Stellenbosch/Bloemfontein cohort (SA)

Odds ratio of increasing HBcrAg with liver disease outcomes accounting for age, HBeAg and HBV VL in cohorts in the UK and SA

Relationship between HBcrAg and other biomarkers in cohorts in the United Kingdom (UK) and South Africa (SA).

Relationship between HBcrAg and thresholds used to assess requirement for perinatal antiviral prophylaxis in UK and SA cohort

## Data Availability

All data produced in the present work are contained in the manuscript or available upon reasonable request to the authors

---

**Suppl Fig 1:**
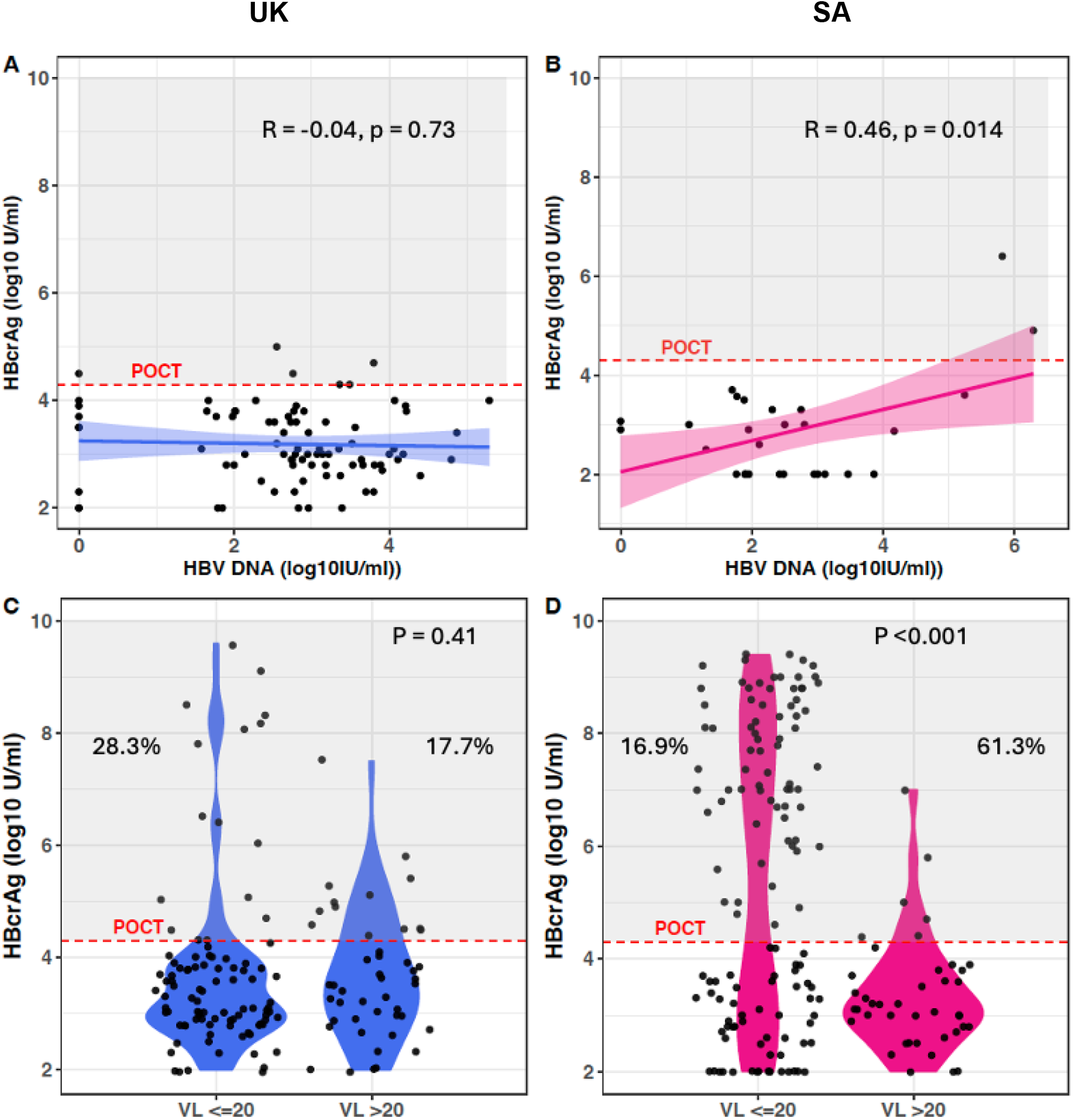
Relationship between HBcrAg and other biomarkers in cohorts in the United Kingdom (UK) and South Africa (SA). A: HBcrAg and HBV VL in HBeAg-negative, untreated UK cohort, B: HBcrAg and HBV VL in HBeAg-negative, untreated SA cohort, C: HBcrAg split by HBV VL ≤ or > 20 in all patients in UK cohort, D: HBcrAg split by HBV VL ≤ or > 20 in all patients in SA cohort. For A and B, Pearsons correlation is shown, for C and D, Mann-Whitney-U test for significance is shown. Dotted red line represents the HBcrAg POCT threshold (HBcrAg ≥4.3 U/ml) with the shaded grey area showing all those points that would test positive. Indicated next to each violin is the percentage of tests which meet this threshold.

**Suppl Fig 2:**
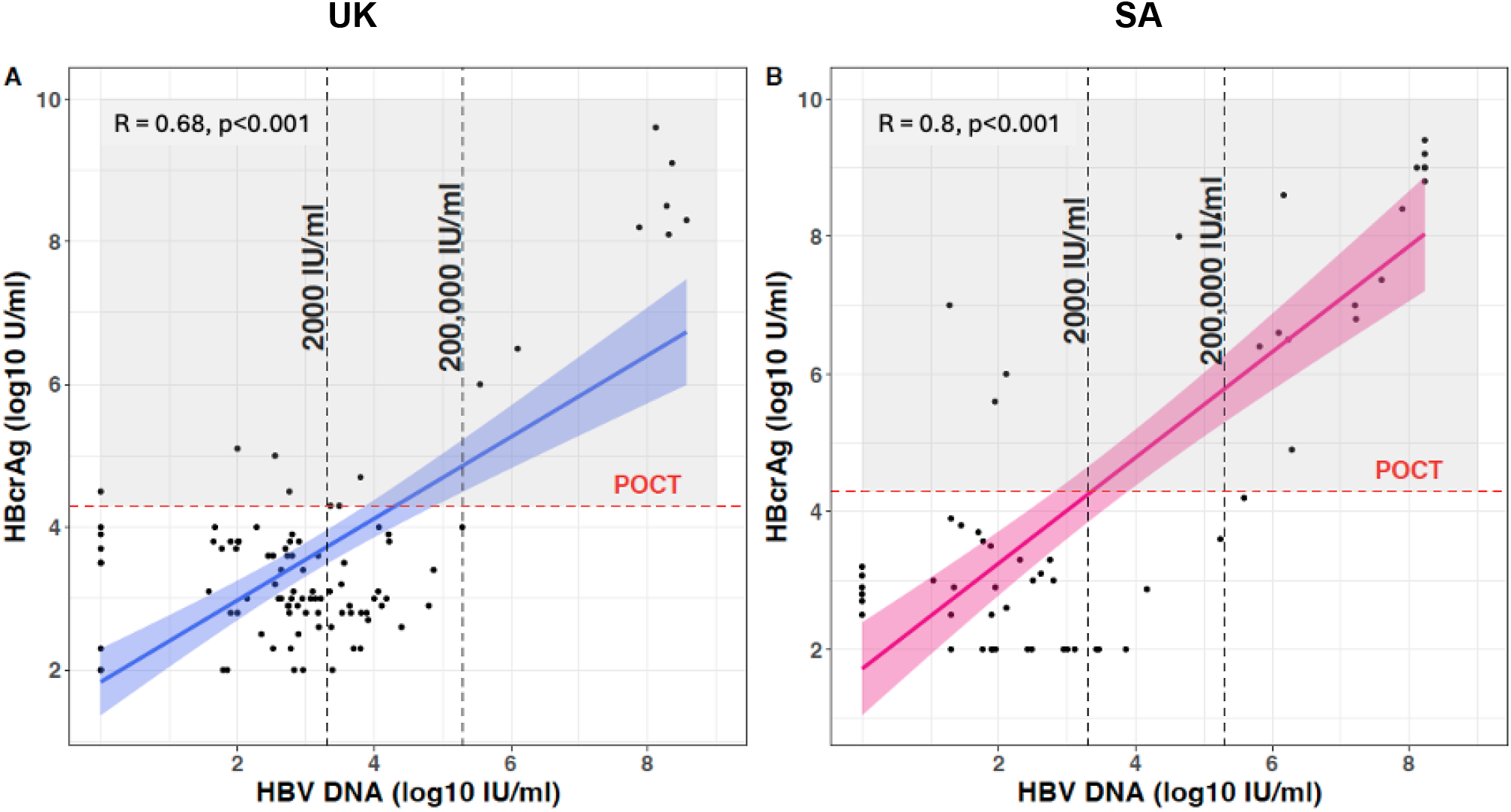
Relationship between HBcrAg and thresholds used to assess requirement for perinatal antiviral prophylaxis in A: UK cohort and B: SA cohort. Data shown for individuals off NA therapy. Vertical black dotted lines show clinically important HBV VL thresholds, and horizontal red dotted lines indicate the HBcrAg POCT threshold. Pearson’s correlation is shown.

**Suppl Table 1:**
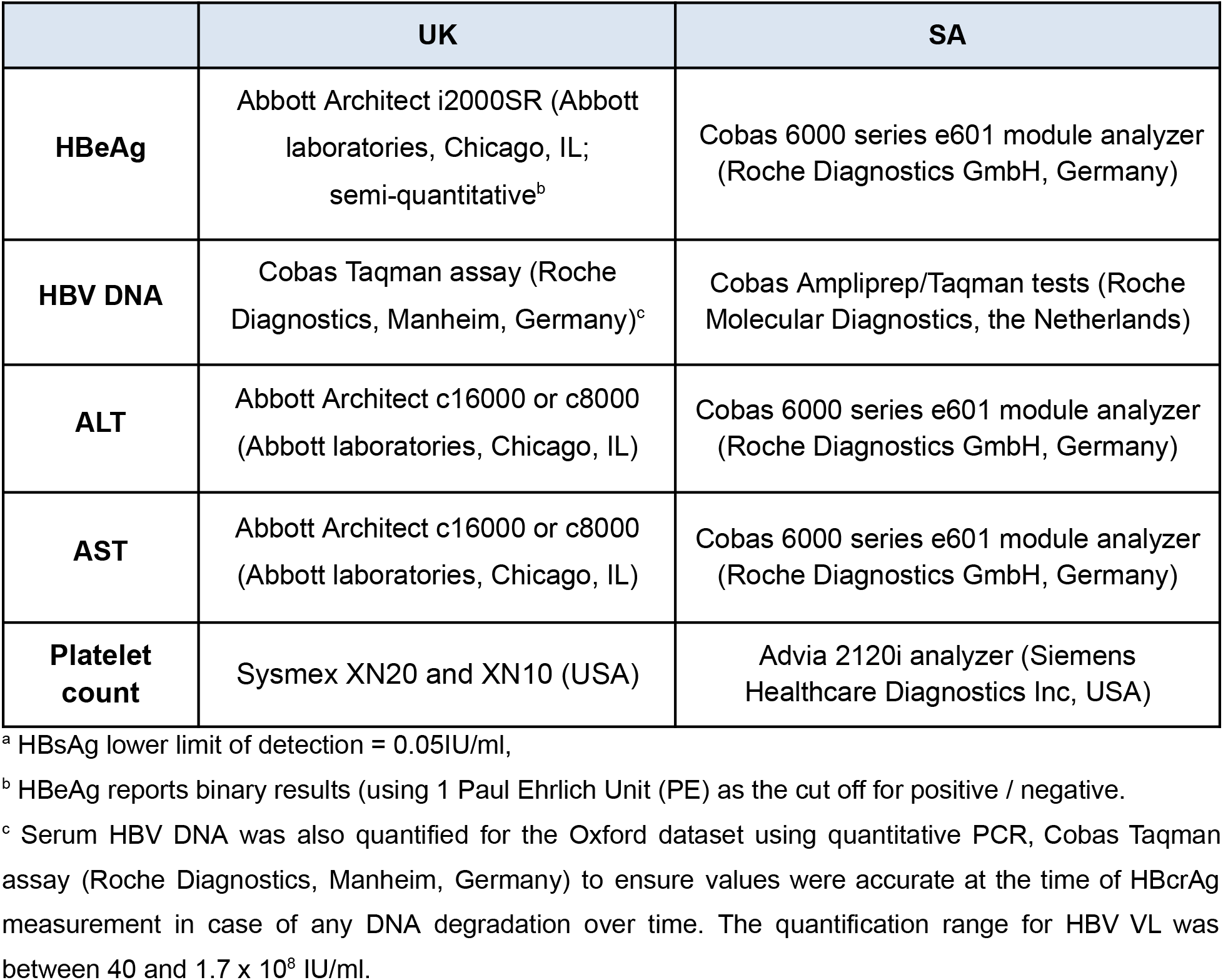
Platforms used to measure routine clinical laboratory biomarkers in Oxford cohort (UK) and Stellenbosch/Bloemfontein cohort (SA)

**Suppl Table 2:**
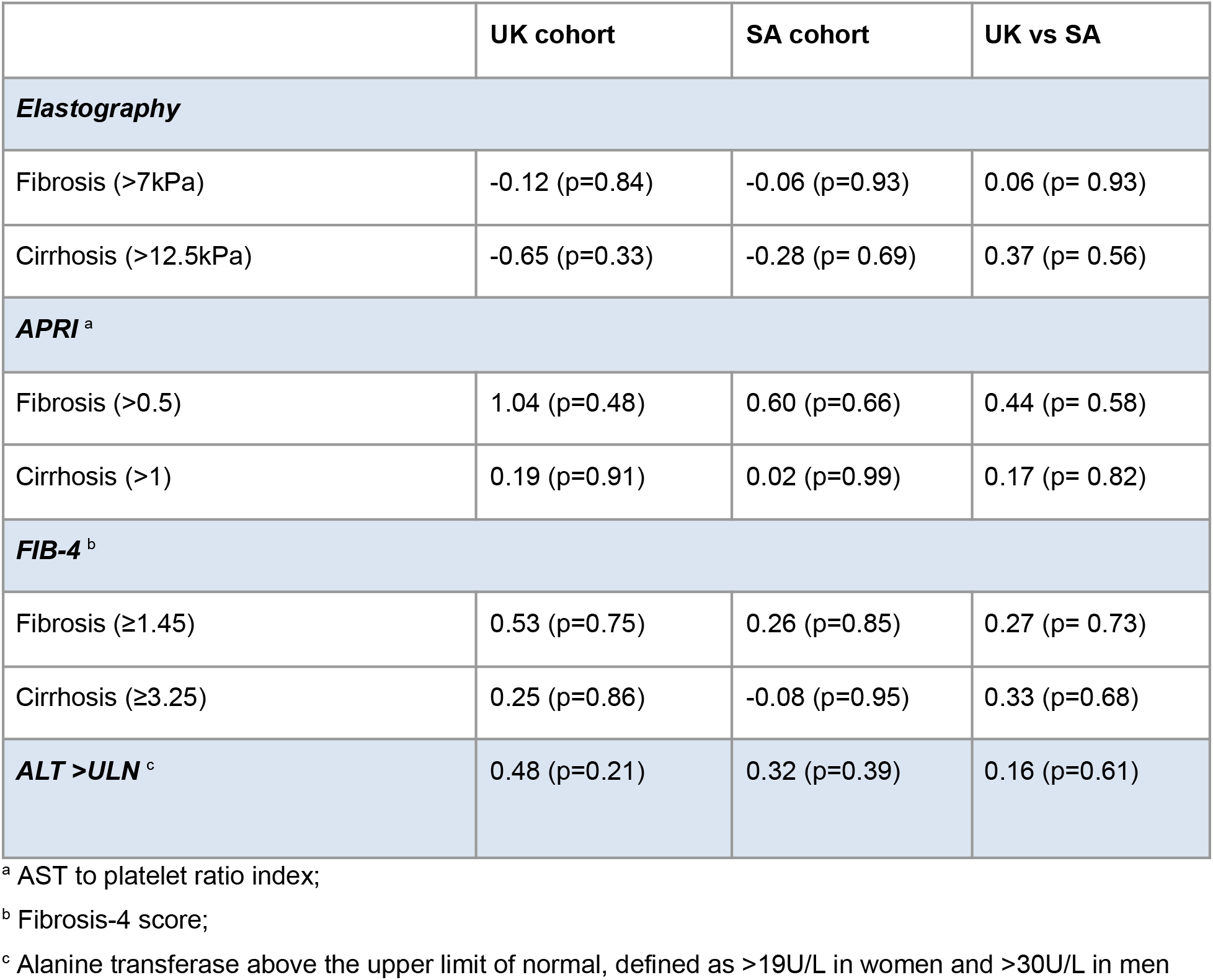
Odds ratio of increasing HBcrAg with liver disease outcomes accounting for age, HBeAg and HBV VL in cohorts in the United Kingdom (UK) and South Africa (SA). Odds ratios (natural logarithm scale) are displayed with associated p-values for each cohort or for the ratio of values between cohorts.

