## Supplementary material for "Evaluation of Hepatitis B core related antigen (HBcrAg) as a biomarker in cohorts from the United Kingdom and South Africa": Platforms used to measure routine clinical laboratory biomarkers in Oxford cohort (UK) and Stellenbosch/Bloemfontein cohort (SA)

**Suppl Table 1: Platforms used to measure routine clinical laboratory biomarkers in Oxford cohort (UK) and Stellenbosch/Bloemfontein cohort (SA)**

| | UK | SA |
|---|---|---|
| **HBeAg** | Abbott Architect i2000SR (Abbott laboratories, Chicago, IL; semi-quantitative[b] | Cobas 6000 series e601 module analyzer (Roche Diagnostics GmbH, Germany) |
| **HBV DNA** | Cobas Taqman assay (Roche Diagnostics, Manheim, Germany)[c] | Cobas Ampliprep/Taqman tests (Roche Molecular Diagnostics, the Netherlands) |
| **ALT** | Abbott Architect c16000 or c8000 (Abbott laboratories, Chicago, IL) | Cobas 6000 series e601 module analyzer (Roche Diagnostics GmbH, Germany) |
| **AST** | Abbott Architect c16000 or c8000 (Abbott laboratories, Chicago, IL) | Cobas 6000 series e601 module analyzer (Roche Diagnostics GmbH, Germany) |
| **Platelet count** | Sysmex XN20 and XN10 (USA) | Advia 2120i analyzer (Siemens Healthcare Diagnostics Inc, USA) |

[a] HBsAg lower limit of detection = 0.05IU/ml,

[b] HBeAg reports binary results (using 1 Paul Ehrlich Unit (PE) as the cut off for positive / negative.

[c] Serum HBV DNA was also quantified for the Oxford dataset using quantitative PCR, Cobas Taqman assay (Roche Diagnostics, Manheim, Germany) to ensure values were accurate at the time of HBcrAg measurement in case of any DNA degradation over time. The quantification range for HBV VL was between 40 and $1.7 \times 10^8$ IU/ml.
