## Supplementary material for "Evaluation of Hepatitis B core related antigen (HBcrAg) as a biomarker in cohorts from the United Kingdom and South Africa": Odds ratio of increasing HBcrAg with liver disease outcomes accounting for age, HBeAg and HBV VL in cohorts in the UK and SA

**Suppl Table 2: Odds ratio of increasing HBcrAg with liver disease outcomes accounting for age, HBeAg and HBV VL in cohorts in the United Kingdom (UK) and South Africa (SA).** Odds ratios (natural logarithm scale) are displayed with associated p-values for each cohort or for the ratio of values between cohorts.

| | UK cohort | SA cohort | UK vs SA |
|---|---|---|---|
| ***Elastography*** | | | |
| Fibrosis (>7kPa) | -0.12 (p=0.84) | -0.06 (p=0.93) | 0.06 (p= 0.93) |
| Cirrhosis (>12.5kPa) | -0.65 (p=0.33) | -0.28 (p= 0.69) | 0.37 (p= 0.56) |
| ***APRI*** [a] | | | |
| Fibrosis (>0.5) | 1.04 (p=0.48) | 0.60 (p=0.66) | 0.44 (p= 0.58) |
| Cirrhosis (>1) | 0.19 (p=0.91) | 0.02 (p=0.99) | 0.17 (p= 0.82) |
| ***FIB-4*** [b] | | | |
| Fibrosis (≥1.45) | 0.53 (p=0.75) | 0.26 (p=0.85) | 0.27 (p= 0.73) |
| Cirrhosis (≥3.25) | 0.25 (p=0.86) | -0.08 (p=0.95) | 0.33 (p=0.68) |
| ***ALT >ULN*** [c] | 0.48 (p=0.21) | 0.32 (p=0.39) | 0.16 (p=0.61) |

[a] AST to platelet ratio index;

[b] Fibrosis-4 score;

[c] Alanine transferase above the upper limit of normal, defined as >19U/L in women and >30U/L in men
