## Supplementary material for "Evaluation of Hepatitis B core related antigen (HBcrAg) as a biomarker in cohorts from the United Kingdom and South Africa": Relationship between HBcrAg and other biomarkers in cohorts in the United Kingdom (UK) and South Africa (SA).

**Suppl Fig 1: Relationship between HBcrAg and other biomarkers in cohorts in the United Kingdom (UK) and South Africa (SA).** A: HBcrAg and HBV VL in HBeAg-negative, untreated UK cohort, B: HBcrAg and HBV VL in HBeAg-negative, untreated SA cohort, C: HBcrAg split by HBV VL ≤ or > 20 in all patients in UK cohort, D: HBcrAg split by HBV VL ≤ or > 20 in all patients in SA cohort. For A and B, Pearsons correlation is shown, for C and D, Mann-Whitney-U test for significance is shown. Dotted red line represents the HBcrAg POCT threshold (HBcrAg ≥4.3 U/ml) with the shaded grey area showing all those points that would test positive. Indicated next to each violin is the percentage of tests which meet this threshold.

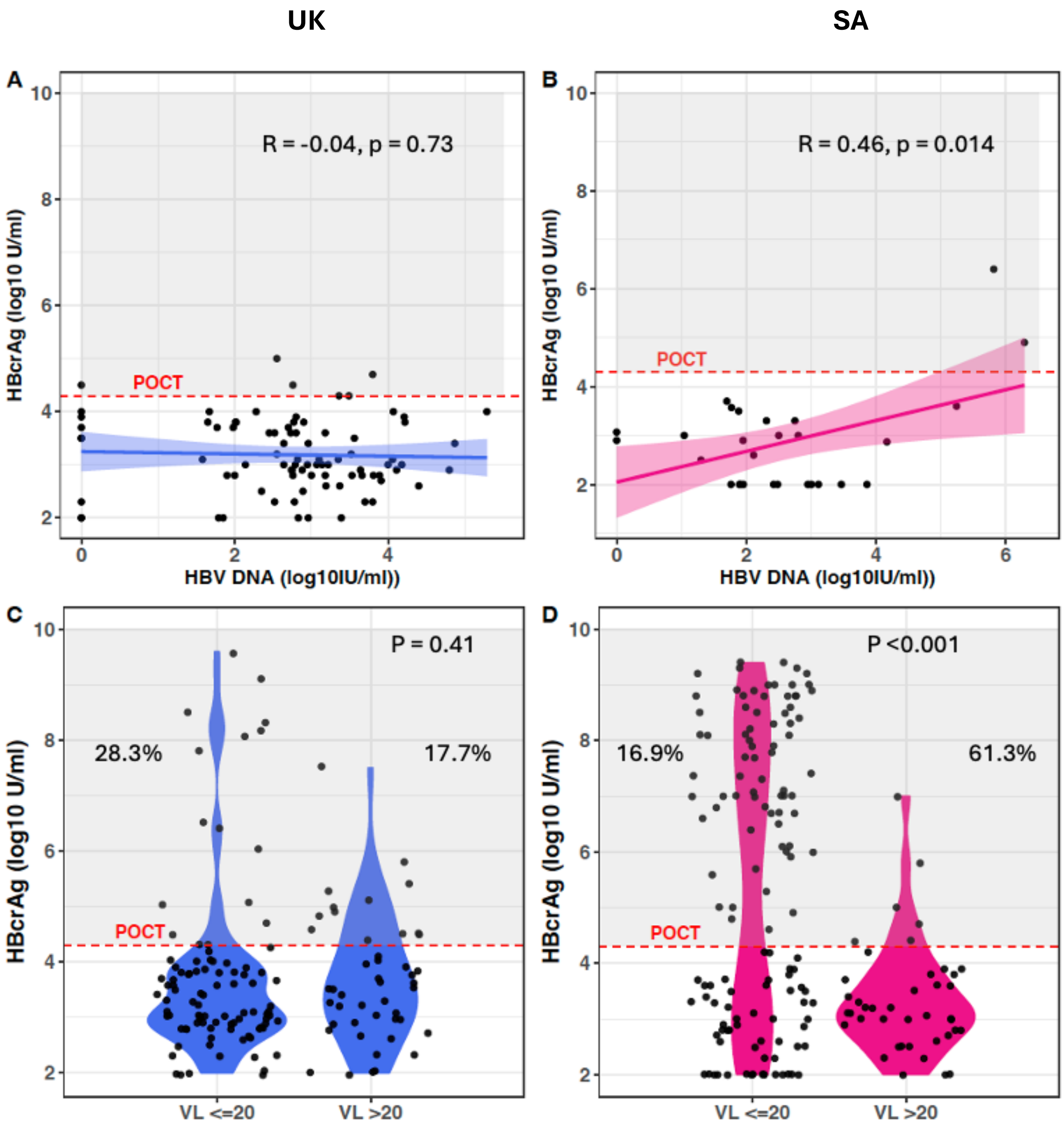
