## Supplementary material for "Evaluation of Hepatitis B core related antigen (HBcrAg) as a biomarker in cohorts from the United Kingdom and South Africa": Relationship between HBcrAg and thresholds used to assess requirement for perinatal antiviral prophylaxis in UK and SA cohort

**Suppl Fig 2: Relationship between HBcrAg and thresholds used to assess requirement for perinatal antiviral prophylaxis in A: UK cohort and B: SA cohort.** Data shown for individuals off NA therapy. Vertical black dotted lines show clinically important HBV VL thresholds, and horizontal red dotted lines indicate the HBcrAg POCT threshold. Pearson's correlation is shown.

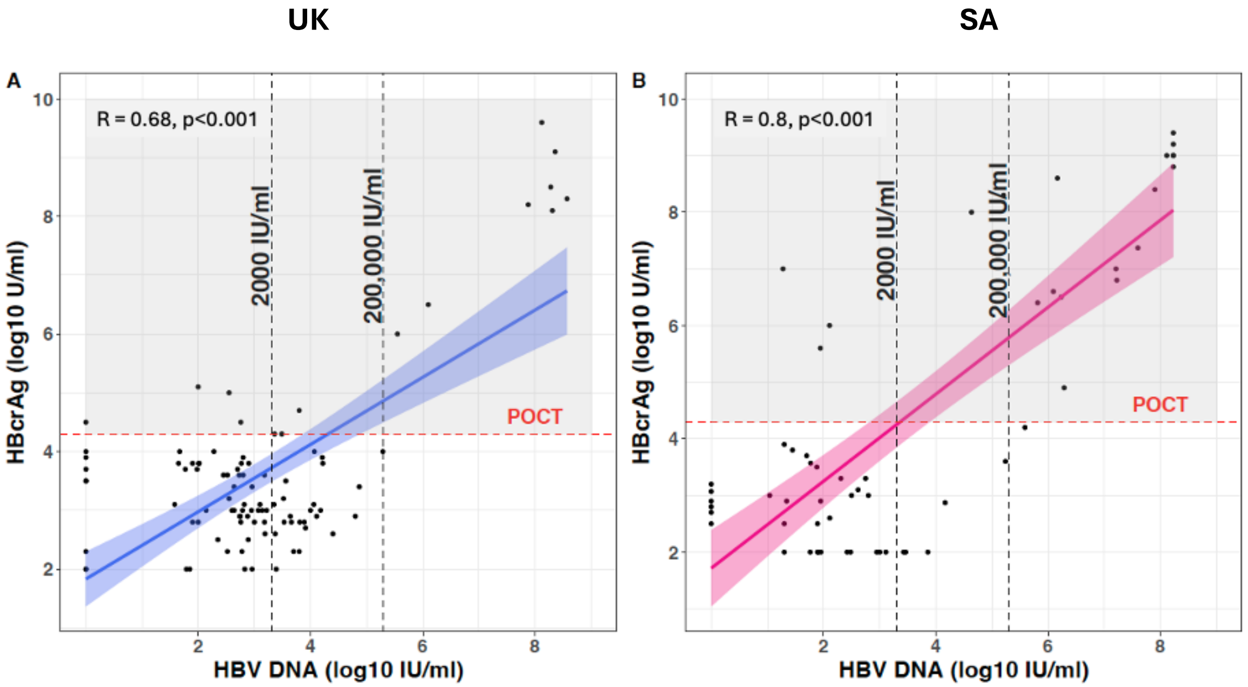
